# Utilizing Metabolomics to Identify Potential Biomarkers and Perturbed Metabolic Pathways in Osteoarthritis: A Systematic Review

**DOI:** 10.1101/2022.09.13.22279898

**Authors:** Peyton Van Pevenage, Jaedyn T. Birchmier, Ronald K. June

## Abstract

**Purpose:** Osteoarthritis (OA) is a joint disease that is clinically diagnosed using components of history, physical exam, and radiographic evidence of joint space narrowing. Currently, there are no laboratory findings that are specific to a diagnosis of OA. The purpose of this systematic review is to evaluate the state of current studies of metabolomic biomarkers that can aid in the diagnosis and treatment of OA.

**Methods:** Articles were gathered from PubMed and Web of Science using the search terms “osteoarthritis” and “biomarkers” and “metabolomics”. Last search of databases took place July 20^th^, 2021. Duplicates were manually screened, along with any other results that were not original journal articles. Only original reports involving populations with diagnosed primary or secondary OA (human participants) or surgically induced OA (animal participants) and a healthy control group for comparison were considered for inclusion. Metabolites and metabolic pathways reported in included articles were then manually extracted and evaluated for significance based on reported *a priori* p-values and/or area under the receiver-operator curve (AUC).

**Results:** Of the 129 results that were returned in the database searches, 36 unique articles met the inclusion criteria. Articles were categorized based on body fluid analyzed: 6 studies on urine samples, 12 studies on plasma samples, 9 studies on synovial fluid (SF) samples, 8 studies on serum samples, and 1 study that involved both plasma and synovial fluid. To synthesize results, individual metabolites, as well as metabolic pathways that involve frequently reported metabolites, are presented for each study. Indications as to whether metabolite levels were increased or decreased are also included if this data was included in the original articles.

**Conclusions:** These studies clearly show that there are a wide range of metabolic pathways perturbed in OA. For this period, there was no consensus on a single metabolite, or panel of metabolites, that would be clinically useful in early diagnosis of OA or distinguishing OA from a healthy control. However, many common metabolic pathways were identified in the studies, including TCA cycle, fatty acid metabolism, amino acid metabolism (notably BCAA metabolism and tryptophan metabolism via kynurenine pathway), nucleotide metabolism, urea cycle, cartilage metabolism, and phospholipid metabolism. Future research is needed to define effective clinical biomarkers of osteoarthritis from metabolomic and other data.

## Introduction

Osteoarthritis (OA) is the most prevalent form of arthritis and has profound detrimental impacts on the patient population burdened by this disease. Approximately 54 million adults in the US have been diagnosed with some form of arthritis, with about 33 million of these being diagnosed with OA.^1^ Factors such as age, obesity, and injury, among many others, are involved in the development of OA; as the US population ages and rates of obesity are on the rise, the prevalence of OA is expected to increase as the total number of arthritis cases, which includes OA diagnoses, is predicted to rise to 78 million US adults by the year 2040.^1^ As OA negatively affects activity levels and quality of life for patients, it can be more difficult to control chronic conditions that OA patients may be concurrently facing, such as diabetes, hypertension, or mental health disorders. OA is also associated with an increased lifetime risk of developing heart disease by 50% as well as an increased all-cause mortality of 55%.^1,2^ As there are no treatments or therapies currently FDA approved and available that can reverse the progression of OA, it is important to make the diagnosis early so that changes can be made to slow the progression and limit the associated negative impacts on quality of life and overall health of patients. Metabolomic profiling is an emerging technology that provides systemic insight into disease processes by characterizing large numbers of biochemical reactants and products, termed metabolite features. As such, there is great potential for using metabolomics to support improved and early diagnosis of OA. Development of metabolomic biomarkers of OA has further potential to identify novel targets for both therapeutic intervention and monitoring disease progression.

OA has historically been considered a degenerative disease, and its development and progression originally were attributed to “wear and tear”.^3^ However, in recent decades our understanding of OA development has evolved to include chronic low-grade inflammation in addition to maladaptive signaling that activate a cascade of catabolic processes across multiple tissues in the joint.^4^ Ultimately in OA the homeostatic balance between maintenance and degradation is upset resulting in deterioration of bone and cartilage.^4^ However the metabolic links between disrupted homeostasis and the pathogenesis of OA are not completely understood, and are an emerging topic of active research utilizing metabolomic analysis.^3-5^ Metabolomics is a growing research field that characterizes large numbers of small-molecules using advanced technology. Metabolomic profiling provides insight into the myriad biochemical reactions that occur in cells and tissues, and pathologically-altered metabolites have the potential to serve as molecular biomarkers of disease processes. The objective of this review is to build upon prior knowledge^6^ and address the current state of metabolomic biomarkers for OA.

The scope of this review includes metabolomic findings by H^1^-NMR, various mass spectroscopy techniques, and immunosorbent assays related to primary and secondary OA. By performing a systematic review involving individual metabolites associated with OA and metabolic pathways that are perturbed in OA disease pathogenesis, this review will summarize both insight into OA provided by metabolomics and candidate biomarkers for future studies. The insight provided here may support and guide further research to enable earlier diagnosis of OA using metabolite biomarkers.

## Methods

The aim of this study was to perform a systematic review of the current state of published metabolomic attempts to discover biomarkers of osteoarthritis.^7^ A search of PubMed and Web of Science was performed and included the search terms: “metabolomics” and “osteoarthritis” and “biomarkers”. A table depicting the search terms for each database is given below in **Table 1**. Included articles were published up until July 20^th^, 2021, with no specific start date filter. To ensure a broad range of studies, there were no filters used for species or language, although all results were in the English language. Initial results were then manually reviewed by PVP to detect duplications. After duplicate articles were removed, the remaining articles were independently reviewed by PVP to remove any results that were not original refereed journal articles (*e*.*g*. conference proceedings) or did not meet the inclusion and exclusion criteria stated below. Reasons for excluding articles were noted and discussed between PVP and RKJ until a consensus was reached. The articles remaining to be included were then grouped based on body fluid utilized in the study: 12 studies of plasma, 6 studies of urine, 9 studies of synovial fluid (SF), 8 studies of serum, and one study of both SF and plasma.

**Table 1.**
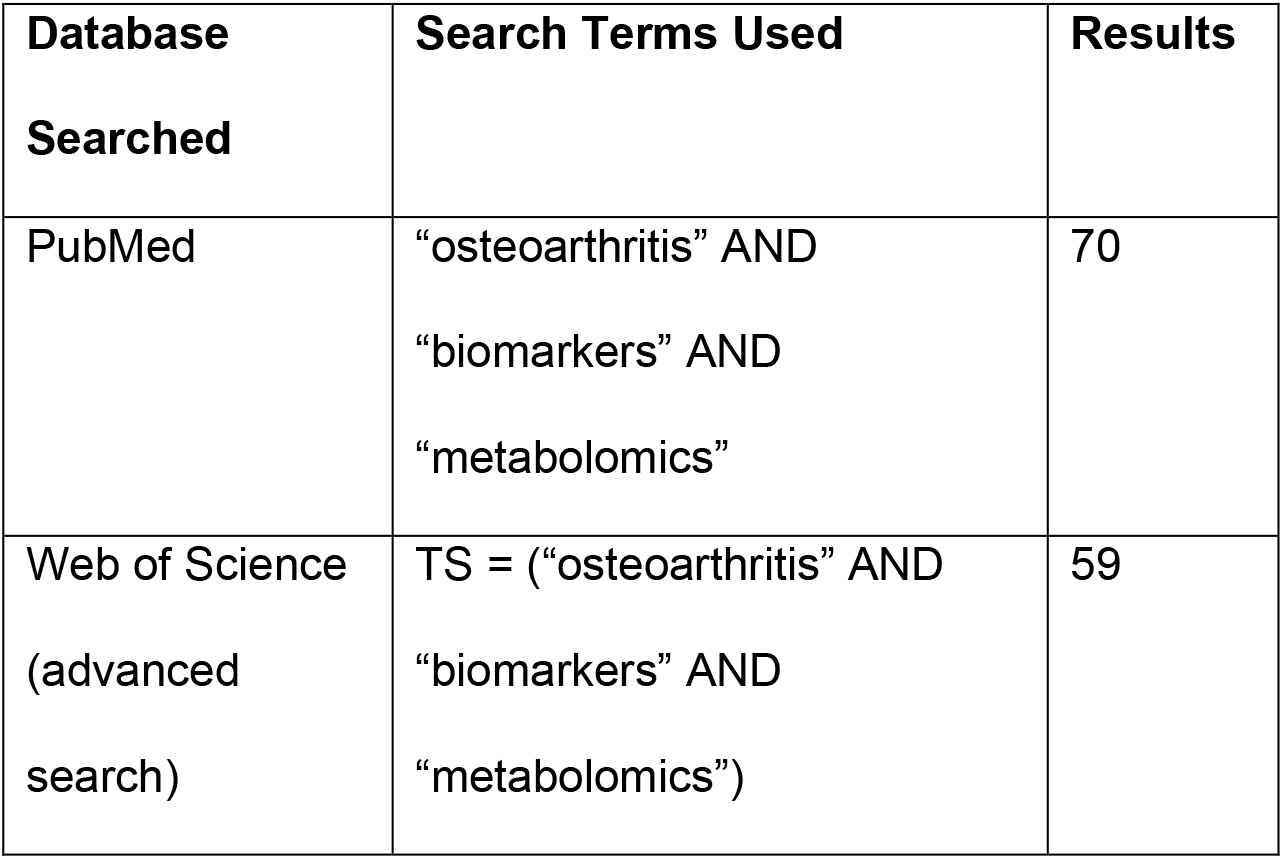
Search Strategy including search terms used for identifying articles in each database.

| **Database Searched** | **Search Terms Used** | **Results** |
| --- | --- | --- |
| PubMed | “osteoarthritis” AND<br>“biomarkers” AND<br>“metabolomics” | 70 |
| Web of Science<br>(advanced search) | TS = (“osteoarthritis” AND<br>“biomarkers” AND<br>“metabolomics”) | 59 |

### Inclusion and Exclusion Criteria

Only original journal articles were included in this review; reviews, abstracts, and brief reports were screened out. Articles were considered for inclusion only if the study population included a group with diagnosed primary or secondary OA (for studies of human participants) or surgically induced OA (for non-human subjects) and a healthy control to allow for comparison. Publications were excluded if they (a) evaluated metabolites specific to therapeutic interventions but did not identify baseline metabolomic profiles or (b) did not identify potential biomarkers of OA with statistical significance. For inclusion, articles also needed to include data describing the metabolomic baseline of healthy controls, or the metabolomics involved in osteoarthritic joints. Additional exclusion criteria included (a) evaluation of animal OA with a distinct pathogenesis differing from that of humans (i.e., OA in equine athletes arising specifically due to carpal osteochondral fragmentation). Articles also needed to include both measures of statistical significance for metabolites and predictive diagnostic value, including p-values or area under the receiver-operator curve (AUC). For metabolite features or pathways to be deemed significant they needed to reach an *a priori* significance threshold of p < 0.05 or AUC > 0.8.

### Data Abstraction

After abstracts were screened using the inclusion and exclusion criteria mentioned, papers considered for inclusion were read in their entirety. Key metabolite features and metabolic pathways were manually extracted based on p-values below the *a priori* threshold of 0.05 or AUCs greater than 0.80. Additional data that was extracted from the articles included the name of first author, year published, body fluid sampled, method of metabolite analysis, and number of subjects in the groups studied. Metabolites that were identified as significant were then categorized according to the particular body fluid that was studied, and metabolites were further grouped based on metabolic pathway involved and biochemical classification. Key metabolic pathways were also categorized according to the body fluid sampled. Data extraction was performed independently by PVP and then reviewed with RKJ.

### Result Synthesis

Eligibility for inclusion in data synthesis was determined based on if significant *a priori* or AUC values were included in the studies. Data from each study was manually extracted and transferred into a Microsoft Excel. The data was then further synthesized by grouping key metabolites based on biochemical class metabolic pathway for each fluid type sampled. This allowed for evaluation of metabolic pathways that were perturbed and offers insight into pathogenesis of osteoarthritis as a disease. No methods were needed for data conversion or handling of missing summary statistics, as each study included *a priori* and/or AUC values for metabolites presented.

Tables were used to present the extracted metabolite and metabolic pathway. For each body fluid studied, a table was created; significant metabolite and metabolic pathway data extracted was then populated in a row in the table corresponding to the fluid evaluated in the study. Grouping of results based on the fluid that was studied allowed for identification of replicated metabolites, as well as common metabolic pathways identified.

### Bias Evaluation

Bias was analyzed at the study and fluid sample level. All disclaimers and statements of competing interest were evaluated. Patient recruitment/selection processes were evaluated; information, or lack thereof, on concurrent health conditions of patients were taken into consideration; targeted versus untargeted metabolomic analysis strategies were noted; finally, methods of acquiring fluid samples (*e*.*g*. clinical patient samples vs cadaver samples) were all taken into consideration. All results reported should be evaluated with the consideration that there are comorbidities that can confound systemic metabolomic attempts to identify biomarkers specific to OA in unknown ways, as the pathophysiologic link between OA and many health conditions have not been discovered or fully investigated.

## Results

Using the search terms described above, 129 total articles were gathered from PubMed and Web of Science. After applying inclusion and exclusion criteria, 36 articles remained for this review. 46 excluded papers were duplicates between the search results of the two databases. 28 excluded results were not original articles and instead consisted of brief reports, reviews, abstracts, editorial material, and a textbook chapter. The 55 remaining articles were then analyzed to determine inclusion: 19 of these were excluded that were either not relevant to OA metabolomics or possible biomarkers, intended to evaluate therapeutic effects of herbal medicine for treatment of OA, evaluated metabolomics to predict responses to total joint replacements in OA, or analyzed metabolomics of OA in a species and disease model that was specific to the species and therefore not generalizable to OA in humans. The remaining 36 articles were included in the review as they offered insight into metabolomics of OA versus healthy controls (34 articles) or provided data on baseline healthy serum (1 article) and plasma (1 article) amino acid levels for reference (**Figure 1**).

**Figure 1:**
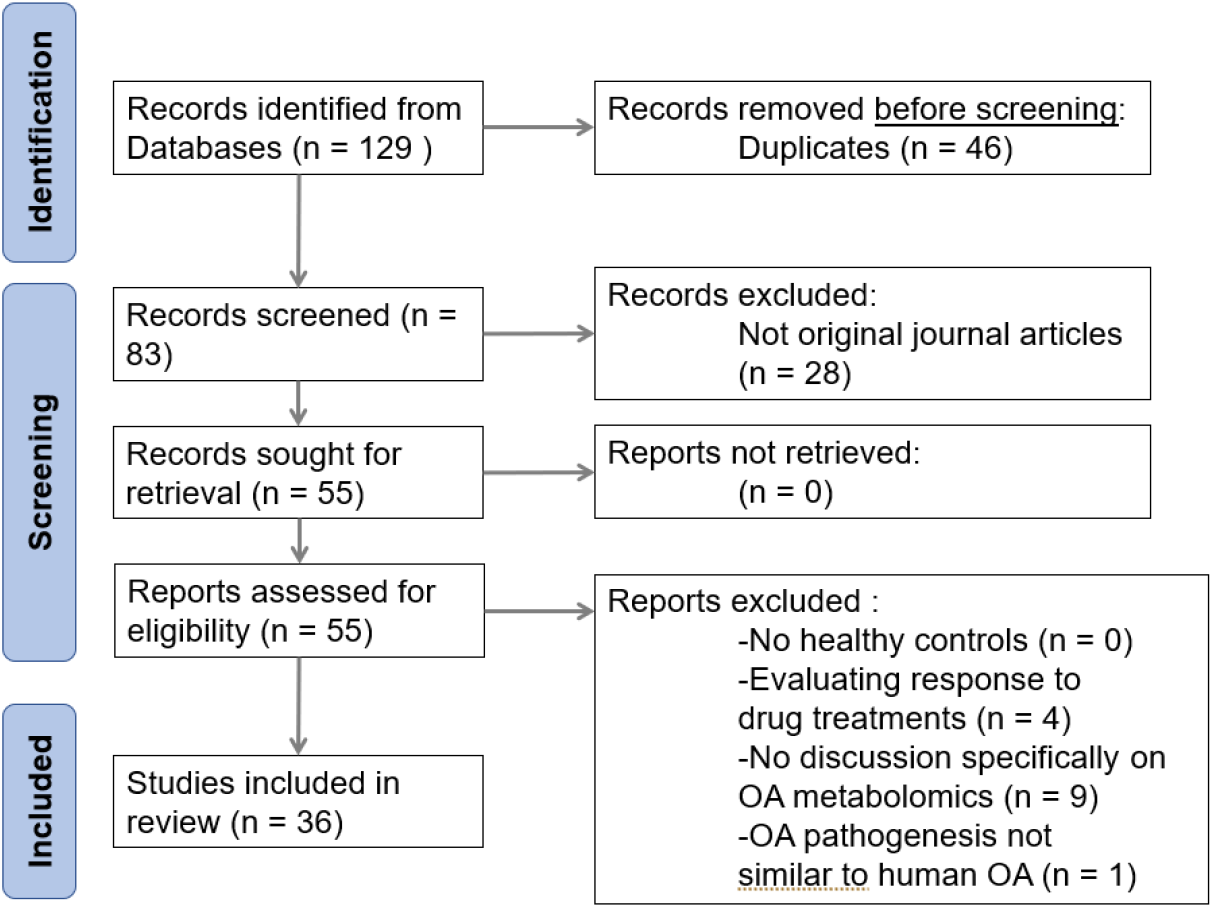
Flow diagram of literature search performed on July 20, 2021, displaying the study selection process.

### Study Characteristics

Upon initial review of the articles, general characteristics of each study were extracted (**Table 2**). These data included the article author, specific body fluid that was used for metabolomic profiling, the model species, and the sample sizes of the various groups. This allowed for further categorization of metabolomic results into the fluids that were analyzed in the studies: plasma, urine, synovial fluid, and serum.

**Table 2.** Study characteristics

| **Article Author** | **Fluid Sampled** | **Model Species** | **Fluid Analysis Method** | **Sample size(s)** |
| --- | --- | --- | --- | --- |
| **Rockel et al., 2018<sup>8</sup>** | Plasma | Human | LC-MS/MS | OA=152, HC=194 |
| **Steventon et al., 2012<sup>9</sup>** | Plasma | Human | RP-HPLC with fluorescence detection | HC=100 |
| **Smilde et al., 2005<sup>10</sup>** | Urine | Guinea Pig | H<sup>1</sup> NMR | OA=12, HC=6 |
| **Carlson et al., 2018<sup>11</sup>** | Synovial Fluid | Human | LC-MS | OA=5, HC=5 |
| **Loeser et al.,**<br>**2016<sup>12</sup>** | Urine | Human | H<sup>1</sup> NMR, ELISA,<br>automated<br>immunoanalyzer | OA1=22, OA2=22 |
| **Carlson et al.,**<br>**2019<sup>13</sup>** | Synovial<br>Fluid | Human | LC-MS | OA GI-II=55, OA GIII-IV=17, HC=7 |
| **Thompson et**<br>**al., 2011<sup>14</sup>** | Serum | Human | FI-ESI-MS | OA=40 |
| **Costello et al.,**<br>**2020<sup>15</sup>** | Plasma | Human | UPLC-MS, FI-ESI-MS | OA=704 |
| **Hu et al.,**<br>**2020<sup>16</sup>** | Synovial<br>Fluid | Rabbit | UPLC-MS | OA=12, HC=12 |
| **Kang et al.,**<br>**2015<sup>17</sup>** | Synovial<br>Fluid | Human | UPLC-QTOF-MS | OA=10 |
| **Jiang et al.,**<br>**2018<sup>18</sup>** | Urine | Rat | GC-TOF-MS | OA=8, HC=8 |
| **Maher et al.,**<br>**2012<sup>19</sup>** | Serum | Ovine | H<sup>1</sup> NMR | OAm=24, HC=12 |
| **Datta et al.,**<br>**2017<sup>20</sup>** | Plasma | Mouse | RP-LC-MS-MS | n=undefined |
| **Adams et al.,**<br>**2014<sup>21</sup>** | Synovial<br>Fluid | Human | LC-MS, GC-MS,<br>ELISA | OA=20, HC=20 |
| **Huang et al.,**<br>**2020<sup>22</sup>** | Plasma | Human | GC-QTOF-MS | OA=12, HC=20 |
| **Senol et al.,**<br>**2019<sup>23</sup>** | Serum | Human | LC-QTOF-MS-MS | OKOA=14, NOKOA=14,<br>HC=15 |
| **Pousinis et al.,**<br>**2020<sup>24</sup>** | Plasma | Mouse | UHPLC-HR-MS | OA=8, HC=7 |
| **Zhang et al.,**<br>**2016<sup>25</sup>** | Plasma | Human | TQ-UPLC-MS | DC-TKR=64, DC-NTKR=45,<br>RC-TKR=72, RC-NTKR=76,<br>LC-TKR=36, LC-NTKR=122 |
| **Tootsi et al.,**<br>**2018<sup>26</sup>** | Serum | Human | UPLC-MS | OA=70, HC=82 |
| **Mickiewicz et al., 2015<sup>27</sup>** | Synovial<br>Fluid | Human | H<sup>1</sup> NMR, GC-MS | OA=55, HC=13 |
| **Mickiewicz et al., 2015<sup>28</sup>** | Synovial<br>Fluid | Ovine | H<sup>1</sup> NMR | OA=9, HC=9 |
| **Abdelrazig et al., 2021<sup>29</sup>** | Urine | Human | LC-HR-MS | OA=74, HC=68 |
| **Liu et al.,**<br>**2021<sup>30</sup>** | Plasma | Human | UPLC-MS, FI-ESI-MS | n=610 |
| **Yin et al.,**<br>**2017<sup>31</sup>** | Urine | Rat | GC-MS | OA=8, HC=8 |
| **Zhang et al.,**<br>**2016<sup>32</sup>** | Plasma | Human | TQ-UPLC-MS | DC-OA=64, DC-HC=45,<br>RC-OA=72,<br>RC-HC=76 |
| **Zhang et al.,**<br>**2016<sup>33</sup>** | Synovial<br>Fluid,<br>Plasma | Human | TQ-UPLC-MS | DC-OA=54, DC-HC=30,<br>RC-OA=72,<br>RC-HC=71 |
| **Maerz et al.,**<br>**2018<sup>34</sup>** | Serum | Rat | H<sup>1</sup> NMR, LC-MS-MS | OA=18, HC=18 |
| **Hu et al.,**<br>**2016<sup>35</sup>** | Plasma | Human | TQ-UPLC-MS | DC-OA=64, DC-HC=45,<br>RC-OA=72,<br>RC-HC=76 |
| **Chen et al.,**<br>**2018<sup>36</sup>** | Serum | Human | TQ-UPLC-MS | OA=32, HC=35 |
| **Zhai et al.,**<br>**2021<sup>37</sup>** | Plasma | Human | TQ-UPLC-MS | OAup=57, OAnup=206,<br>OAbp=177, OAnbp=116 |
| **Zhai et al.,**<br>**2010<sup>38</sup>** | Serum | Human | LC-MS-MS | DC-OA=123, DC-HC=299,<br>RC-OA=76, RC-HC=100 |
| **Zhang et al.,**<br>**2015<sup>39</sup>** | Serum | Human | UPLC-MS | OA=40, HC=20 |
| **Kosinska et**<br>**al., 2014<sup>40</sup>** | Synovial<br>Fluid | Human | FI-ESI-MS-MS, LC-MS-MS | eOA=17, IOA=13, HC=9 |
| **Hügler et al.,**<br>**2012<sup>41</sup>** | Synovial<br>Fluid | Human | H<sup>1</sup> NMR | OA=15 |
| **He et al.,**<br>**2021<sup>42</sup>** | Plasma | Human | GC-MS | OA=12, HC=12 |
| **Li et al., 2009<sup>43</sup>** | Urine | Human | GC-MS | OA=37, HC=37 |

**Table 2**: General characteristics of each study include first author and date of publication, fluid sampled, model species, method of fluid analysis for metabolomics, and sample sizes. LC=Liquid chromatography, MS=Mass spectrometry, RP=Reversed phase, HPLC=High-performance liquid chromatography, H^1^-NMR=Proton Nuclear Magnetic Resonance, ELISA=Enzyme-linked immunosorbent assay, FI-ESI=Flow-injection electrospray ionization, QTOF=quadrupole time-of-flight, GC=Gas chromatography, TOF=Time-of-flight, TQ=Triple quadrupole, HR=High resolution, UPLC=Ultra performance liquid chromatography, OA=Osteoarthritis subjects, HC=Healthy control for comparison, OA1=Evidence of OA progression on radiographic findings 18 months from baseline, OA2=No evidence of OA progression on radiographic findings 18 months from baseline, OA GI-II=OA grades I-II using Outerbridge scale scoring, OA GIII-IV=OA grades III-IV using Outerbridge scale scoring, OAm=Models with OA induced by meniscal destabilization (MD, 12) or anterior cruciate ligament transection (ACLT, 12), OKOA=Knee OA and obesity (BMI≥30), NOKOA=Knee OA without obesity (BMI<30), TKR=Total knee replacement, NTKR=No total knee replacement performed, DC=Discovery cohort subjects, RC=Replication cohort subjects, LC=Longitudinal study cohort subjects, OAup=OA patients with radiographic evidence of unilateral knee OA progression, OAnup=OA patients without radiographic evidence of unilateral knee OA progression, OAbp=OA patients with radiographic evidence of bilateral knee OA progression, OAnbp=OA patients without evidence of bilateral knee OA progression, eOA=OA with Outerbridge classification scale score ≤ 2, lOA=OA with Outerbridge classification scale score >2.

### Risk of Bias in Studies

Bias was difficult to assess. In human studies, patient ethnicities were only reported in 2 of 36 articles.^12,37^ Potentially confounding concurrent health conditions were usually not determined or reported. Since obesity accompanies ∼90% of knee OA cases, hypertension accompanies about 40% of knee OA cases, and diabetes accompanies about 16% of knee OA cases^2^ this may impact patient metabolomic profiles. For selection of human control groups, patients were usually selected on the basis that they lacked a diagnosis of OA by a family medicine provider and/or lacked radiographic evidence of OA. However subclinical or asymptomatic forms of OA may be present in these HC groups, and this could potentially confound metabolomic comparisons. For human cases where body fluids were taken from live patients, controls were also matched for age, sex, and BMI to limit demographic variation between groups.

In animal models where OA was surgically induced, controls underwent sham surgeries in attempts to ensure that differences in metabolite profiles were a reflection of OA and not surgical complications (e.g. incision-related inflammation). The final source of bias occurs at the study level and involves the use of either targeted or untargeted metabolomics: both have limitations for analyzing the comprehensive body of metabolites present in a body fluid due to the large numbers of potential metabolites.

### Synthesis of Results

For analysis, studies were grouped by body fluid; metabolites reported with statistical significance were extracted along with identified metabolic pathways, and are presented below.

### Urine

Six studies evaluated metabolites found in urine, significant metabolites are summarized in **Table 3** below; three of these studies used humans as subjects,^12,29,43^ two of the studies used Sprague-Dawley rats,^18,31^ and one study used the Hartley outbread guinea pig model that spontaneously develops progressive knee OA starting at ∼10 months of age^10^. No single metabolite was identified to be significantly altered across all 6 of the urine metabolite studies. Increased alanine concentrations in OA versus HC were identified in two studies with p-values <0.01.^18,31^ Histidine was identified to be significantly altered in three studies;^12,31,43^ two studies reported a statistically significant decrease in histidine concentrations between OA and HC with p-values <0.05.^31,43^ Histidine levels were 1.3x higher (p-value 0.016) in OA patients with BMI ≥27kg/m^2^ who had radiographic progression of knee OA at 18 months versus OA patients with BMI ≥27kg/m^2^ who did not have radiographic progression of knee OA at 18 months^12^ (Radiographic progression was defined as decrease in minimum joint space width on radiograph ≥0.7mm compared to baseline radiographs 18 months prior). Glutamine was found to be significantly decreased in three studies with p-value <0.05.^18,31,43^

**Table 3:**
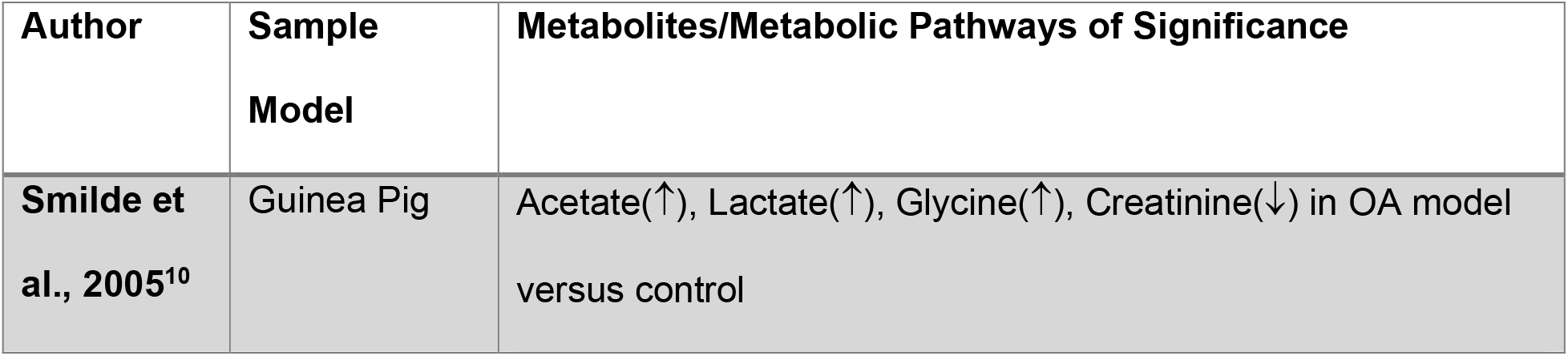
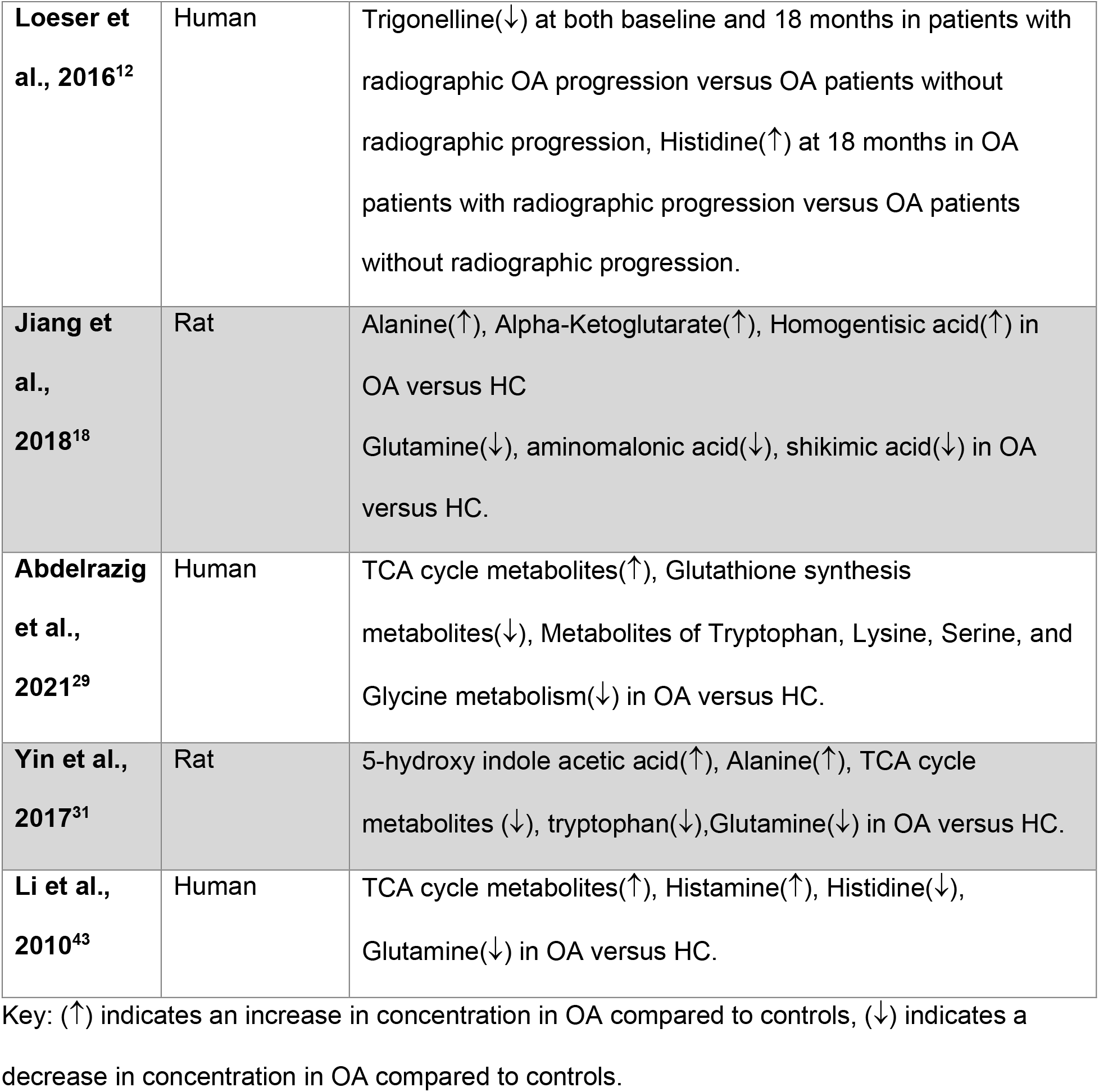
Significant Metabolites Found in Urine

The remaining significantly altered metabolites detected in the studies of urine mapped to many metabolic pathways. These include amino acid metabolism (phenylalanine, tryptophan, alanine, histidine, lysine, serine, threonine, glycine, glutamine, asparagine), lipid metabolism, nucleotide metabolism, the TCA cycle, ATP storage and utilization, and oxidative stress.

### Plasma

13 studies evaluated OA metabolites found in plasma (Table 4). 10 studies evaluated plasma metabolites of OA in humans^8,9,15,22,25,30,32,33,35,37,42^, 2 studies evaluated plasma metabolites of OA in mice^20,24^. One study evaluated baseline plasma amino acid levels and was included for reference of absolute concentrations of plasma amino acids.^9^ Ten of the 12 plasma metabolite studies found lysophosphatidylcholine (LysoPC) and phosphatidylcholine (PC) species to be statistically significantly altered in OA^8,15,20,24,25,30,32,33,35,37^. Among these 10 studies that reported LysoPC and PC metabolism to be altered in OA, Rockel *et al* found LysoPC and PC signatures to be statistically significant only in males over the age of 50 years old compared to HC,^8^ while Liu *et al* found LysoPC and PC levels to be statistically significant only between OA with multi-joint pain versus OA with single joint pain but not OA versus HC.^30^

**Table 4:** Significant Metabolites Found in Plasma

| Author | Sample Model | Metabolites/Metabolic Pathways of Significance in OA vs HC |
| --- | --- | --- |
| Rockel et al., 2018<sup>8</sup> | Human | Lysophosphatidylcholine and phosphatidylcholine species were found to be altered in OA versus healthy, strongest signature in males >50 years of age. |
| Steventon et al., 2012<sup>9</sup> | Human | This study did not report metabolites, and instead was included as a reference as it reported normal plasma amino acid levels. |
| Costello et al., 2020<sup>15</sup> | Human | Glutamine, BCAAs, PCaaC38:0, and PCaaC40:6 found to overlap most between OA patients without function or pain response after undergoing total knee replacement. |
| Datta et al., 2017<sup>20</sup> | Mouse | Lysophosphatidylcholine and phosphatidylcholine levels remained elevated 3 months after returning to a normal chow diet from a high fat diet in OA mice model |
| Huang et al., 2020<sup>22</sup> | Human | Succinic acid(↓), Xanthurenic acid(↓), L-tryptophan(↓) in OA versus HC. |
| Pousinis et al., 2020<sup>24</sup> | Mouse | Cholesteryl ester species(↑), phosphatidylcholine species(↑), Sphingomyelin d34:1(↑) in OA versus HC. |
| Zhang et al., 2016<sup>25</sup> | Human | Ratio of total lysophosphatidylcholines to total phosphatidylcholines was on average 0.05μM higher in OA versus HC. Increased BCAA to histidine ratio seen in OA versus HC. |
| Liu et al., 2021<sup>30</sup> | Human | Lysophosphatidylcholine and phosphatidylcholine metabolism was altered in OA patients with multi-joint pain versus OA |
|  |  | patients with pain in one joint, however not significant when comparing OA versus HC. |
| **Zhang et al., 2016<sup>32</sup>** | Human | Arginine(↓) Lysophosphatidylcholines species(↓), Phosphatidylcholine species(↓), Proline(↑) in OA versus HC. |
| **Zhang et al., 2016<sup>33</sup>** | Human | Altered phosphatidylcholine metabolism with phosphatidylcholine species(↓) was shared between OA without diabetes and OA with diabetes and also found independently in OA and diabetes, although the link between these diseases and pathways is unknown. Separation between OA with MetS and OA with type II diabetes mellitus versus OA without MetS appeared to be due to type II diabetes as opposed to hypertension, dyslipidemia, or obesity in MetS. |
| **Hu et al., 2016<sup>35</sup>** | Human | In differential correlation network, Ac-ornithine, Arginine, Alanine, and ornithine were found to be significant and made up the core of the network. Glycerophospholipids (lysophosphatidylcholine and phosphatidylcholine) species prominent in between the core and periphery of the network. |
| **Zhai et al., 2021<sup>37</sup>** | Human | Phenylalanine was significantly associated with both unilateral and bilateral knee OA progression from baseline to 30-month follow-up, noted to be a better predictor in women. Arginine, Leucine, Isoleucine, and Carnitine species also associated with unilateral progression. Serine, Tryptophan, Histidine, Lysine, Ornithine, Asparagine, and Phosphatidylcholine species were associated with bilateral progression. |
| He et al.,<br>2021<sup>42</sup> | Human | Cholesterol(↑), Stearic acid(↑), L-Lactic acid(↑), Alpha-Tocopherol(↑), Oxalic Acid(↑) in OA versus HC. |

Key: (↑) indicates an increase in concentration in OA compared to controls, (↓) indicates a decrease in concentration in OA compared to controls.

Although some articles evaluated LysoPC and PC levels in OA overall, others found these two classes of molecules to be altered in various conditions and models. In addition to findings of altered plasma LysoPC and PC levels broadly in OA, LysoPC and PC levels were found to be altered in other settings; for example, altered LysoPC and PC levels compared to health control were seen in the context of metabolic changes in OA with high fat diets^20^, differential correlation networks for patient pain and function responses to total joint replacements^15^, progression of OA in either unilateral and/or bilateral knee OA^37^, or in OA associated with metabolic syndrome (MetS) or components of MetS^33^. While LysoPC and PC species were mentioned in these ten plasma metabolomics articles, there was not a clear consensus as to whether LysoPC and PC species were increased or decreased in OA plasma compared to HC.

Amino acid levels were also altered in many studies. However, as with LysoPC and PCs, increases or decreases were not always specified. Glutamine, isoleucine, proline, tryptophan, arginine, alanine, phenylalanine, serine, histidine, lysine, and asparagine were all reported to be significantly altered with p-values <0.05.^15,22,25,32,35,37^

The remaining significantly altered metabolites corresponded largely to altered fatty acid metabolism, TCA cycle, and steroid biosynthesis.

### Synovial Fluid

10 studies analyzed SF. Of these, 8 used a human model^11,13,16,17,21,27,28,41^, 1 used a New Zealand White rabbit model^16^, and 1 study used an ovine model^28^ (**Table 5**). In the synovial fluid studies, many metabolites mapped to metabolic pathways involving fatty acids, phospholipids, amino acids, vitamins, TCA cycle, collagen breakdown, glycosaminoglycan and keratin sulfate degradation, urea cycle, and oxidative stress.

**Table 5:** Significant Metabolites Found in Synovial Fluid

| **Authors** | **Sample Model** | **Metabolites/Metabolic Pathways of Significance in OA vs HC** |
| --- | --- | --- |
| **Carlson et al., 2018<sup>11</sup>** | Human | Phospholipid species, Fatty Acid metabolites, Acylcarnitine species, Ursolic acid and Sulfonic acid derivatives were associated with differentiating OA versus HC. |
| **Carlson et al., 2019<sup>13</sup>** | Human | Altered extracellular matrix component metabolism (glucosamine and galactosamine biosynthesis, ascorbate metabolism, keratin sulfate metabolism, N-glycan metabolism), amino acid metabolism, lipid metabolism (glycosphingolipid, glycerophospholipid, and carnitine/FA metabolism), leukotriene metabolism, central energy (glycolysis, gluconeogenesis, TCA cycle), oxidative stress (vitamin E and glutathione) and vitamin C, E, B1, B3, B6, and B9 metabolism in OA versus HC. |
| **Hu et al., 2020<sup>16</sup>** | Rabbit | Polyamines (acetylspermidine species), Phospholipid species, Tryptophan and Tryptophan metabolites (including kynurenic acid), Acylcarnitine Species, Phenylalanine and Phenylalanine |
|  |  | metabolites, Lysine, Arginine, and Histidine were associated with differentiating OA versus HC. |
| **Kang et al., 2015<sup>17</sup>** | Human | Tryptophan metabolism, arachidonic acid metabolism, acylcarnitine metabolism, phospholipid metabolism, phenylalanine and tyrosine metabolism altered in OA. |
| **Adams et al., 2014<sup>21</sup>** | Human | IL-1Ra(↑), IL-6(↑), IL-8(↑), IL-10(↑), IL-15(↑), MCP-1(↑), Tryptophan(↑), Kynurenine(↑), pro-hydroxy-proline(↑), FA species(↑), Glutamate(>7x↑), Glutathione metabolites(↑), Isoleucine(↑) in post-traumatic ankle arthritis versus HC. |
| **Mickiewicz et al., 2015<sup>27</sup>** | Human | Citrate(↑), Malate(↓), Acylcarnitine species(↓), Creatine(↓) in OA versus HC. |
| **Mickiewicz et al., 2015<sup>28</sup>** | Ovine | Serine(↓), Asparagine(↓), Hydroxyproline(↓), Uridine(↓), Isobutyrate(↑), and Glucose(↑) reported as signs of early degenerative OA changes after surgical ACL-injury induced OA versus sham surgery HC. |
| **Zhang et al., 2016<sup>33</sup>** | Human | Altered phosphatidylcholine metabolism with phosphatidylcholine species(↓) was shared between OA without diabetes and OA with diabetes and also found independently in OA and diabetes, although the link between these diseases and pathways is unknown. Separation between OA with MetS and OA with type II diabetes mellitus versus OA without MetS appeared to be due to type II diabetes as opposed to hypertension, dyslipidemia, or obesity in MetS. |
| **Kosinska et al., 2014<sup>40</sup>** | Human | Sphingomyelin, Ceramide, Hexosylceramides, Phosphatidic Acid, Lysophosphatidic Acid, Phosphatidylglycerol, and Lysylphosphatidylglycerol species all increased in OA versus HC, and also increased between IOA compared to eOA. |
| **Hügler et al., 2012<sup>41</sup>** | Human | Hippuric acid, Phenylalanine, Tyrosine, D-Glucose, Valine, TCA cycle metabolites, Carnitine species, Glutamine, Lactate, Alanine, and Lysine signatures present on H<sup>1</sup> NMR of OA. |

Key: (↑) indicates an increase in concentration in OA compared to controls, (↓) indicates a decrease in concentration in OA compared to controls.

Phospholipid species were not only frequently found to be significantly altered between OA and controls,^11,13,16,17,33,40^ but phospholipid levels also continued to increase when comparing early OA disease (Outerbridge score ≤ 2) to late OA (Outerbridge score >2) and the HC population.^40^

Metabolites indicating altered energy metabolism were also commonly identified in SF of OA compared to HC. These included FA metabolites, TCA cycle metabolites, gluconeogenic amino acids, and sugars in the synovial fluid samples.^11,13,16,17,21,27,28,41^ Multiple carnitine species were perturbed; one study indicated decreased levels of two acylcarnitine species in OA synovial fluid.^27^ Two of the other studies mentioned altered acylcarnitine species as statistically significant OA predictors (*e*.*g*. AUC >0.90), although it was not clear if the particular acylcarnitine species increased or decreased in OA compared to HC.^11,16^ Altered acylcarnitine levels in the SF of OA compared to HC may indicate altered energy production, as carnitine and carnitine-acylcarnitine translocase allow acyl chains to move across the inner mitochondrial membrane and into the mitochondrial matrix where beta-oxidation occurs.^44^

The prominence of altered metabolites that are involved in mitochondrial energy production and metabolism, as well as evidence of oxidative stress, may indicate mitochondrial dysfunction in the osteoarthritic joint.

### Serum

8 studies involving serum were included in this review (**Table 6**): five articles analyzed human serum OA metabolites^23,26,36,38,39^, one article studied serum in rats^34^, one article examined serum in an ovine model^19^, and one article evaluated daily variation of acylcarnitines and amino acids in OA patients without comparison to a HC group^14^. Four of the articles reported altered levels of branched-chain amino acids between OA and HC groups.^19,23,36,38^ Two studies reported statistically significant increases in valine, isoleucine, and leucine.^36,38^ One study reported increased valine levels in OA serum.^23^ The fourth article reported decreases in BCAAs in an ovine model of induced OA via surgical ACL transection.^19^ Maher et al. suggest that their observed decrease in BCAAs may be specific to the ovine species, as literature reporting increased BCAAs present in human OA serum was reported prior to their publication^38^ and has been replicated in other studies,^23,36^ and therefore should be investigated further.^19^

**Table 6:** Significant Metabolites Found in Serum

| Author | Sample Model | Metabolites/Metabolic Pathways of Significance in OA vs HC |
| --- | --- | --- |
| **Thompson et al., 2011<sup>14</sup>** | Human | This study did not report metabolites, and instead was included as a reference as it reported daily variations of serum amino acid and acylcarnitine levels. |
| **Maher et al., 2012<sup>19</sup>** | Ovine | BCAAs(↓), 3-Methylhistidine(↑), Glycine(↑), Creatinine(↑), Creatine(↑) at 4 weeks and 12 weeks in post-ACL Transection OA model versus HC. |
| **Senol et al., 2019<sup>23</sup>** | Human | Phospholipid species(↑), Indoleacetic acid(↑), Urea(↑), Valine(↑), Alanine(↑), FA metabolites(↑) increased in OA versus Control. |
| **Tootsi et al.,**<br>**2018<sup>26</sup>** | Human | Total Acylcarnitine species to Carnitine species ratio was decreased in OA versus HC. Carnitine Palmitoyltransferase 1 activity was decreased in OA versus HC. |
| **Maerz et al.,**<br>**2018<sup>34</sup>** | Rat | Acylcarnitine species increased at 72 hours, 4 weeks, and 10 weeks post-ACL rupture OA model versus HC. Most significantly affected pathways in OA versus HC identified as cyanoamino acid metabolism, methane metabolism, sphingolipid metabolism, histidine metabolism, glutathione metabolism, tryptophan metabolism, beta-alanine metabolism, fructose and mannose metabolism, and amino and nucleotide sugar metabolism. |
| **Chen et al.,**<br>**2018<sup>36</sup>** | Human | BCAAs(↑), Tryptophan(↑), Alanine(↑), 4-Hydroxy-L-Proline(↑), Creatine(↑), Arginine(↑), Lysine(↑), Tyrosine(↑), Glutamine (↓), Phenylalanine(↓), Serine(↓), Proline(↓), Gamma-Aminobutyric Acid(↓), Creatinine(↓), Taurine(↓), Asparagine(↓), Acetyl-Carnitine(↓), Citrulline(↓) in OA versus HC. |
| **Zhai et al.,**<br>**2010<sup>38</sup>** | Human | Increased BCAA to Histidine ratio in OA versus HC, with ratio of Valine to Histidine and Isoleucine to Histidine having strongest statistical significance. |
| **Zhang et al.,**<br>**2015<sup>39</sup>** | Human | CRP(↑), Homocysteine(↑), Tryptophan(↑), Glycine(↓), Histidine(↓) in OA versus HC. |

Key: (↑) indicates an increase in concentration in OA compared to controls, (↓) indicates a decrease in concentration in OA compared to controls.

Acylcarnitine species were serum metabolites associated with OA. One study found a lower ratio of total acylcarnitine to carnitine with lower levels of 8 out of 14 acylcarnitines species\ measured in OA versus HC.^26^ They also found lower levels of carnitine palmitoyltransferase 1 activity.^26^ CPT-1 is present on the outer mitochondrial membrane and is the rate limiting enzyme for long-chain fatty acid beta-oxidation the mitochondrial matrix. Decreased activity of this enzyme may drive mitochondrial dysfunction in OA. Another study noted a decrease in acetyl-carnitine in OA compared to HC.^36^ The fourth serum study to mention acylcarnitine species noted a significant increase in seven acylcarnitine species (p = 0.008-0.041) over 10 weeks after ACL rupture-induced post-traumatic OA.^34^

Other statistically significant serum metabolites mapped to phospholipid metabolism, amino acid metabolism (tryptophan, arginine, proline, serine, alanine, glycine, histidine, tyrosine), TCA cycle, nucleotide metabolism, energy metabolism, and the urea cycle.

## Discussion

Metabolomics is a growing field of research that can offer insight into disease pathophysiology and develop metabolic biomarkers that support early diagnosis and intervention, as well as both monitoring of disease progression and evaluation of treatment efficacy. OA is a disease with emerging metabolic features, and there are currently no treatment options that stop or reverse disease progression.^45^ Therefore, identification and diagnosis of OA early in the disease course is critical to prevent extensive cartilage destruction and allow modifiable risk factors to be addressed to slow disease progression.

The studies included in this review analyzed the metabolomics of OA in urine, serum, plasma, and synovial fluid. The studies were done with humans, rats, mice, sheep, guinea pigs, and rabbits. Various analytical methods including mass spectrometry were performed to identify and compare metabolites between OA and comparison groups in each biological fluid. Metabolites that resulted from analysis of body fluids were either reported as being altered, while other studies further clarified whether the measured concentrations of reported metabolites were increased or decreased in comparison to control groups. In studies that reported increased or decreased concentrations, the data compiled by the authors shows that approximately 168 metabolites were increased in OA versus control out of 239 metabolites. This proportion was more or less consistent across each individual fluid sample.

In Maher et al^19^, the findings of decreased BCAA concentrations in ovine OA subjects is contrary to reported increases in BCAA concentrations in OA in other studies.^23,36,38^ Maher et al suggest that their observation of decreased BCAA concentrations in OA may result from interspecies differences in disease metabolic processes. This highlights the need for further studies of OA metabolomics and pathophysiology, as well as further evaluation of animal models of OA. Identifying both similarities and differences between animal and human OA is important to ensure that metabolomic findings are relatable across species. Biomarker candidates identified in animal models must be evaluated in human clinical populations to ensure their utility.

The heterogeneity of OA is reflected in the reported metabolites and pathways detailed in these studiers. From these studies, there was no consensus on either a single metabolite or a panel of metabolites that could serve as predictive biomarkers for early diagnosis of OA. This reflects the complexity of the disease and the multi-tissue pathophysiology that drive the progression of OA.

Although there was variation in the specific metabolites reported in these studies, common metabolic themes arose among many of the reports. Metabolites associated with mitochondrial dysfunction were detected in several studies. These include alterations in fatty acid metabolism, acylcarnitine and carnitine metabolites, and TCA cycle metabolites. Additionally several studies found metabolites indicating the presence of reactive oxygen species, gluconeogenic amino acid metabolites, and metabolites of antioxidants. There is potential for future studies to expand upon these results by focusing on metabolites associated with mitochondrial dysfunction.

Common themes seen across many studies include metabolism of phospholipids and articular cartilage, as well as metabolism of many amino acids, notably BCAAs. Altered levels of phospholipid metabolites including lysophosphatidylcholine and phosphatidylcholine species, sphingomyelins, ceramides, hexosylceramides, phosphatidylglycerols were noted; levels of proline and hydroxyproline, as well as glutamate which is used by mammals to synthesize proline, were noted in many studies and are major constituents of type II collagen found in articular cartilage. Altered amino acid metabolism was a common finding among many of the articles, with BCAA to histidine ratio being reported as a possible diagnostic marker of OA. Although the changes in the levels of BCAAs and histidine in the studies were not unanimously agreed upon across the studies, it warrants further investigation and research as two studies claimed that BCAA to histidine ratio could be a statistically significant marker of OA. Due to their frequent recurrence across studies, metabolites of phospholipid and articular cartilage breakdown, as well amino acid metabolism, could be topics of further investigation in future studies.

### Tryptophan Metabolism

Tryptophan can be synthesized and degraded through multiple pathways including the serotonin and melatonin pathway, the kynurenine pathway, and the indole pathway. Tryptophan metabolism and metabolite levels are altered in many diseases, ranging from chronic inflammatory diseases, autoimmune disorders, and neurodegenerative diseases.^21^ 17 articles included in this review found tryptophan levels to be significantly altered in OA, and 5 articles suggested that the kynurenine pathway may be involved in OA pathogenesis in some way. Kynurenine is produced from tryptophan by indolamine-2,3-oxidase, and this kynurenine production is enhanced after treatment by either interferon gamma or tumor necrosis factor-alpha.^46,47^ Metabolites further downstream in the kynurenine pathway can lead to increased production of reactive oxygen species, as well as modulation of the inflammatory T-cell response to induce tolerance to mild inflammatory attack.^46^ Further study of the tryptophan-kynurenine axis in OA may yield important information for better understanding OA pathogenesis.

### Limitations

36 articles met the inclusion criteria for this review. However, there exists literature on the topic of metabolomics of OA that did not meet our inclusion criteria, or possibly were not displayed in the results of the database searches due to keyword or database limitations, and therefore were not included in this review. As such this review focused on identifying basic metabolite features altered in OA, analyzing how metabolomic profiles change during OA disease progression, and finding relationships between OA and other health conditions like metabolic syndrome, orthopaedic trauma, and cardiovascular disease. The reviewed articles did not always report data on these other health conditions which limits the generalizability of these findings. Additionally, Zhang *et al*., 2016, reported metabolite ratios that were only significant in advanced knee OA but not in early-stage OA.^25^ This suggests that metabolite biomarkers of OA may be stage-specific which suggests their potential to detect progression of OA.

Among all articles in this review, more than 350 metabolites were reported to be altered between OA and control subjects. These metabolites achieved varying degrees of significance with p-values smaller than 0.01 to barely distinguishable statistical differences between OA and HC. While some of these statistically significant metabolite findings were replicated across many studies,, such as LysoPC and PC species^8,11,15,20,23-25,32,33,35,37^ and the amino acids glycine,^10,19,29,39^ alanine,^18,23,31,34-36,41^ and histidine,^12,16,25,34,37-39,43^ and many others, a large majority of individual metabolites were not. However, because metabolic pathways involve detection of subsets of their individual metabolites, comparing reported metabolic pathways was more feasible than detailing individual metabolites in many cases. Thus, it is possible that future studies may identify additional biomarkers based on the pathways reported herein.

As plasma, urine, and serum are all fluids that reflect systemic metabolism, future studies should identify and control for confounding factors, such as concurrent medical conditions, patient demographics, and dietary habits. These key covariates may drive metabolomic changes that require additional statistical modeling for developing biomarkers of OA. Clinical sampling of SF for metabolomics involves inherent risks as it is an invasive procedure. This challenge renders obtaining HC samples challenging. For future studies of OA synovial fluid, it would be beneficial to obtain healthy age- and sex-matched SF samples to allow for direct comparison to OA SF. Because SF directly contacts multiple tissues of the diseased joint, in contrast to systemic fluids, it contains great potential for developing biomarkers of OA and OA progression.

## Conclusions

OA is a heterogenous disease with a wide range of altered metabolic pathways and processes that result in disease pathogenesis and progression. These studies demonstrate this heterogeneity through the vast array of metabolites and pathways reported. Through this review, metabolites indicated abnormalities in fatty acid metabolism and the TCA cycle suggesting mitochondrial dysfunction, altered energy metabolism, amino acid metabolism, nucleotide metabolism, phospholipid metabolism, urea cycle, and cartilage metabolism.

Many reports find differences in lysophosphatidylcholine/phosphatidylcholine metabolism, tryptophan and kynurenine metabolism, and branched-chain amino acid metabolism in OA. These metabolites and pathways have potential both for developing prognostic biomarkers of OA and for improved understanding of OA pathogenesis and progression. Sub-groups and endotypes of OA were also proposed in several reports. Future studies might characterize metabolomic differences between phenotypes and endotypes of OA. The identification of distinct phenotypes or endotypes may support development of novel therapeutic targets and help address modifiable risk factors involved in OA development and progression.

While metabolomic profiling is relatively new to the OA field, future primary and replication studies have good potential to provide reliable, sensitive, and specific panels of biomarkers to allow for early diagnosis of OA.

## Data Availability

This is a systematic review. The references include all of the original reports on which this review is based.

## Acknowledgements

The authors are grateful for funding support from NIH (NIAMS 5R01AR073964) and NSF (1554708 and 2140127).

